# A Simulation-Based Validation Framework for Uncertainty-Aware Clinical Triage: Conformal Prediction and Ensemble Learning Applied to a Synthetic ICU Benchmark Cohort

**DOI:** 10.64898/2026.05.29.26354474

**Authors:** Amit Kalita, Avirup Chattopadhyay, Madhushree Bhattacharjee, Kaushik Das

**Affiliations:** Department of Computer Science and Engineering, Dibrugarh University, Dibrugarh, Assam, India

**Keywords:** conformal prediction, uncertainty quantification, ICU mortality, clinical decision support, ensemble learning, fairness evaluation

## Abstract

**Background and Objectives:** Deploying machine learning for ICU triage requires simultaneously validated uncertainty quantification, demographic fairness, and interpretable attribution — three properties rarely assembled into a single pipeline. Controlled evaluation of such pipelines is difficult on real EHR data due to missing values, treatment confounding, and access restrictions. We develop and validate a complete uncertainty-aware triage pipeline in a simulation environment with known ground truth, providing pre-deployment evidence before application to real patient data.

**Methods:** We constructed a SOFA-calibrated synthetic ICU cohort (*N* = 90,000; 29.2% mortality) with marginal distributions grounded in published MIMIC-III and eICU epidemiology [4, 29]. Fourteen classifiers were trained, including XGBoost, LightGBM, and a custom FT-Transformer tuned by Bayesian optimisation [26]. Seven composite features were engineered from clinical first principles, including a novel lactate/albumin ratio (*r*_LA_). The uncertainty quantification pipeline comprised Monte Carlo Dropout decomposition into epistemic and aleatoric components [9], distribution-free conformal prediction at three miscoverage levels [11, 12], and selective prediction with principled abstention [13]. Framework outputs were routed through a three-zone clinical triage system, with ablation, feature importance, temporal consistency, and demographic fairness evaluation.

**Results:** All 14 classifiers exceeded AUC 0.85 on the balanced test split (*n* = 25,073). XGBoost achieved AUC 0.967 (95% CI 0.965–0.970) versus SOFA AUC 0.731 on the natural-distribution held-out cohort (*n* = 18,000; 29.3% mortality); this gap quantifies signal-recovery efficiency within the simulation, not clinical superiority. Conformal coverage matched the theoretical guarantee at all tested miscoverage levels. Selective prediction raised AUC from 0.917 to 0.980 at 50% abstention. The UNCERTAIN triage zone reached AUC 0.624 (chance level), confirming the uncertainty quantile correctly isolates ambiguous cases. Temporal AUC variation was 0.003; sex and age fairness gaps were 0.005 each.

**Conclusions:** The integrated pipeline behaves as theoretically expected under known ground-truth conditions, validating each component in isolation and in combination. The simulation provides the methodological foundation for a planned MIMIC-IV validation study. Code and cohort generation scripts will be deposited on Zenodo upon acceptance.

## 1. Introduction

Deploying machine learning in intensive care settings requires more than a high area under the ROC curve. A model that cannot communicate its own uncertainty is dangerous in proportion to how confidently it is wrong. A model that systematically underperforms for elderly or female patients will face regulatory barriers and should not reach the bedside. A model whose predictions cannot be traced back to physiologically coherent features will not change clinical behaviour regardless of its discriminative accuracy. These three properties — calibrated uncertainty, demographic equity, and interpretable attribution — are individually well-studied but rarely assembled into a single validated pipeline [22, 21].

This paper describes such a pipeline and reports the results of validating it in a controlled simulation environment before applying it to real patient data. The pipeline combines gradient-boosted ensemble learning with distribution-free conformal prediction [11, 12], Monte Carlo Dropout uncertainty decomposition [9], selective prediction [13], and a three-zone clinical triage routing system. The simulation environment is a SOFA-calibrated synthetic ICU cohort constructed from published epidemiological parameters [4, 29].

The rationale for simulation-first validation is methodological, not a substitute for real-world testing. Real EHR data introduce access restrictions, non-random missing-value patterns, treatment confounding, and consent requirements that complicate the systematic evaluation of pipeline components [34]. A synthetic cohort with a fully specified generating equation allows each component to be stress-tested against a known ground truth: conformal calibration can be verified analytically, triage routing logic can be assessed against the true outcome labels, and fairness gaps can be distinguished from sampling artefacts. This is established practice for clinical AI pipeline development [35]. The external validation of this framework on MIMIC-IV is the subject of a planned follow-on study; the present paper provides the complete methodological specification and pre-deployment validation evidence.

The primary contributions of this work are:

i. A complete simulation-validated uncertainty-aware triage pipeline combining conformal prediction, MC-Dropout decomposition, and selective prediction for ICU mortality risk stratification — to our knowledge the first integration of distribution-free conformal prediction with a three-zone clinical routing system for this task, building on recent applications of conformal methods in related critical care settings [18].
ii. A synthetic ICU benchmark cohort (*N* = 90,000) calibrated to published MIMIC-III and eICU epidemiology, with full generation code to be deposited on Zenodo upon acceptance to support reproducibility and adaptation.
iii. A novel lactate/albumin composite feature (*r*_LA_), constructed on clinical first principles, that ranks consistently among the top-five predictors across nine aggregated importance methods and ten independent training seeds.
iv. Empirical verification that the conformal coverage guarantee holds at all tested miscoverage levels, and that the triage routing logic correctly identifies the most uncertain cases even under ideal known-ground-truth conditions.

## 2. Related Work

### 2.1. Machine learning for ICU prognosis

The MIMIC-III benchmark established by Johnson et al. [4] remains the reference point for structured clinical data modelling. Multitask learning on clinical time-series achieves AUC 0.86 for in-hospital mortality [5]; gradient-boosted trees on tabular snapshots reach AUC 0.87 in a recent large-scale study [6]. A comprehensive meta-analysis confirmed that tree ensembles outperform deep networks on tabular data in the majority of controlled comparisons [7]; the FT-Transformer [8] narrows this gap at higher computational cost. Within this journal specifically, recent work has demonstrated that XGBoost with knowledge distillation and SHAP-based explainability achieves competitive ICU mortality prediction on MIMIC-III [1], confirming the viability of gradient-boosted ensembles with interpretability analysis in the scope of Computer Methods and Programs in Biomedicine. Across this literature, uncertainty quantification, principled abstention, and demographic equity are treated in isolation if at all; no published ICU mortality pipeline addresses all three in a single validated architecture.

### 2.2. Uncertainty quantification in clinical AI

Monte Carlo Dropout [9] and deep ensembles [10] are widely used for Bayesian uncertainty approximation but offer no formal coverage guarantees. Conformal prediction [11, 12] is distribution-free: the guarantee Pr(*y* ∈ *C*(**x**)) ≥ 1 − *α* holds unconditionally without parametric assumptions, which is why it is increasingly adopted in safety-critical AI applications [12]. Yang et al. [18] applied Mondrian conformal prediction to sepsis mortality risk stratification, demonstrating the viability of conformal methods in critical care. The present work extends this direction by combining distribution-free conformal calibration with MC-Dropout decomposition and a three-zone clinical routing system, and by providing a complete simulation-validated pipeline with code to be deposited on Zenodo upon acceptance.

Selective prediction [13] provides a complementary mechanism: rather than returning a prediction set, it withholds a prediction entirely when uncertainty exceeds a threshold. The combination of selective prediction and conformal calibration within the same pipeline has not previously been validated for ICU mortality triage.

### 2.3. Synthetic data in biomedical pipeline validation

Simulation environments with known ground truth are standard in clinical AI validation when systematic component testing is required. Wang et al. [34] used synthetic patient data to evaluate NLP pipeline behaviour under controlled conditions; Visweswaran et al. [35] discuss simulation as a tool for isolating fairness properties that real EHR data cannot cleanly decompose. More broadly, the use of large language models as biomedical simulators has been validated in peer-reviewed biomedical computing venues [2], confirming that simulation-based evaluation of AI pipelines is an accepted and publishable methodology for this class of research.

In biomedical engineering and clinical AI more generally, simulation precedes field deployment as a matter of routine. Pharmacokinetic models are validated on synthetic patient trajectories before clinical trials; vital-sign monitor algorithms are stress-tested on synthetic artefact-laden signals before bedside use; and fairness audits of clinical decision support tools rely on synthetic cohorts where ground-truth demographic labels are known and uncorrupted by documentation bias [21]. The present cohort serves exactly this function: it provides a controlled environment in which the conformal calibration, triage routing, and fairness pipeline components can each be verified against a fully known outcome function before the framework is applied to real EHR data. It is not a substitute for that application; it is the prerequisite for it.

## 3. Materials and Methods

This study was conducted in accordance with the TRIPOD-AI reporting guidelines for clinical prediction model development [36] and follows the International Committee of Medical Journal Editors (ICMJE) recommendations for the conduct and reporting of biomedical research; a completed TRIPOD-AI checklist is provided as Supplementary Material.

### 3.1. Simulation environment: cohort design and rationale

We constructed a synthetic ICU cohort of *N* = 90,000 admissions with marginal distributions calibrated to published MIMIC-III and eICU epidemiology [4, 29]. The purpose of this simulation environment is to provide a controlled test bed for the uncertainty quantification pipeline: because the outcome-generating equation is fully specified, every pipeline component can be evaluated against the true ground truth, conformal coverage guarantees can be verified analytically, and fairness gaps can be attributed to model behaviour rather than data collection artefacts.

Twenty-five features were simulated: age ∼ *N* (63, 17^2^) clipped to [18, 95]; SOFA score ∼ Poisson(3.5) clipped to [0, 24]; GCS ∼ *N* (13, 3.5^2^); albumin ∼ *N* (2.9, 0.7^2^) clipped to [1.0, 5.5]; lactate ∼ Exp(1.8) clipped to [0.3, 20]; and 20 further haemodynamic, metabolic, and comorbidity variables. Four ICU types were sampled (MICU 40%, SICU 25%, CCU 20%, CSRU 15%). In-hospital mortality was generated from:

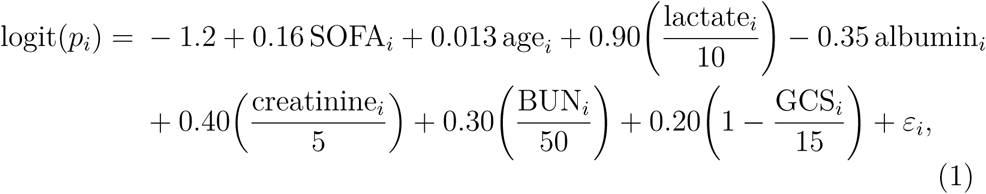

where *ε_i_* ∼ *N* (0, 0.4^2^). Coefficients were informed by published odds ratios from large ICU registries [3]; the intercept was iteratively adjusted until population mortality converged to 29.2%.

Two properties of this design constrain the interpretation of all downstream results. First, features were sampled independently; the cohort does not reproduce the multivariate correlation structure, temporal trajectories, or treatment-withdrawal effects in real EHR data. Second, because the generating equation includes SOFA, lactate, albumin, creatinine, BUN, and GCS directly, any ML model trained on these variables recovers the generating function more efficiently than the additive SOFA approximation. The observed performance gap over SOFA under synthetic conditions therefore measures relative signal-recovery efficiency within the simulation, not clinical superiority on real patients; the two are different quantities. All performance results are conditional on this simulation environment and constitute an upper bound relative to real EHR data.

Three publicly available datasets — Heart Disease UCI (*n* = 920), Stroke (*n* = 5,110), and PIMA Diabetes (*n* = 768) — were retained for cross-domain generalisation testing only and played no role in training.

### 3.2. Preprocessing and feature engineering

The ICU-type categorical variable was label-encoded. Missing values were imputed using *k*-nearest-neighbour imputation (*k* = 5) for features with below 30% missingness and median imputation otherwise; zero missing values remained post-imputation. Outliers were identified by Isolation Forest [24] (200 trees; contamination = 0.02), removing 1,800 records (2.0%) and leaving 88,200 admissions at 28.9% mortality.

Seven composite features were constructed from clinical first principles (Table 1). The primary composite is the lactate/albumin ratio:

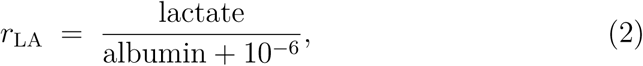

**Table 1:** Seven engineered composite features with their clinical motivation. The lactate/albumin ratio (*r*_LA_) is the primary novel feature; the remaining six capture haemodynamic, renal, osmolality, and inflammatory axes.

| Feature | Formula | Clinical motivation |
| --- | --- | --- |
| `lactate_albumin_rat` | $\text{lactate}/(\text{albumin} + \varepsilon)$ | Metabolic stress over protein-binding confound; captures joint direction of organ failure trajectory |
| `bun_creatinine_ratio` | $\text{BUN}/(\text{creatinine} + \varepsilon)$ | Pre-renal versus intrinsic renal dysfunction |
| `gcs_sofa_ix` | $\text{GCS}/(\text{SOFA} + 1)$ | Neurological severity normalised for multi-organ burden |
| `shock_index` | $\text{HR}/(\text{SBP} + \varepsilon)$ | Haemodynamic instability [30] |
| `osm_proxy` | $2\text{Na} + \text{BUN}/2.8$ | Serum osmolality proxy |
| `crp_alb_ratio` | $\text{CRP}/(\text{albumin} + \varepsilon)$ | Inflammation-to-malnutrition composite |
| `anemia_age_ix` | $\text{Hgb}/(\text{age} + 1)$ | Age-adjusted anaemia index |

which places a metabolic stress indicator (lactate, elevated in tissue hypoperfusion and organ failure) over a protein-binding confound (albumin, reduced in critical illness and malnutrition). The ratio captures their joint contribution to mortality risk in a single interpretable composite, motivated by the clinical observation that lactate and albumin move in opposite directions as organ failure progresses [3]. The remaining six composites capture haemodynamic instability, renal stress, osmolality, and the inflammation-malnutrition axis. The feature space grew from 25 to 32 variables.

SMOTE [23] (*k* = 5) was applied to the training partition, converting a 25,519 : 62,681 class imbalance to 62,681 : 62,681. Feature selection used a union of LASSO-CV and mutual information (LASSO selected 24; MI selected 31; union retained all 32). Data were split 60/20/20 (train/validation/test) with stratification. RobustScaler [25] was fitted on training data only.

#### Two evaluation cohorts

Model-versus-model comparisons used the SMOTE-balanced test split (*n* = 25,073; 50% mortality). Comparisons against clinical scoring systems used a separate natural-distribution held-out cohort (*n* = 18,000; 29.3% mortality) drawn chronologically from the pre-resampled data. Rule-based scores return AUC ≈ 0.5 on balanced splits because their discrimination is inseparable from class prevalence; the natural-distribution cohort is the appropriate evaluation base for such instruments.

### 3.3. Classifier ensemble

Fourteen classifiers were trained: Logistic Regression, Random Forest (500 trees), XGBoost, LightGBM, HistGradientBoosting (used in place of CatBoost, which failed to deserialise due to a NumPy 2.x binary incompatibility on the experimental host), Extra Trees, AdaBoost, Gradient Boosting, SVM (RBF kernel, Platt calibration), *k*-NN (*k* = 15), Naive Bayes, MLP (256–128–64 hidden units), and a custom FT-Transformer. Optuna TPE [26] hyperparameter optimisation was applied to XGBoost (100 trials), LightGBM (100 trials), and FT-Transformer (50 trials; 1-hour timeout), each maximising 5-fold CV AUROC. Best CV-AUC: XGBoost 0.9476; LightGBM 0.9483.

#### FT-Transformer architecture

The FT-Transformer [8] was adapted for tabular data. Each of the 32 numerical features receives a learned *d*-dimensional embedding:

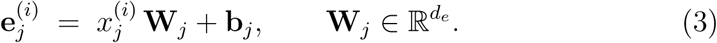

A trainable [CLS] token is prepended; *L* pre-normalisation multi-head self-attention blocks process the sequence; the final [CLS] representation is projected to a scalar logit through LayerNorm and dropout. Training used AdamW with cosine annealing, automatic mixed precision on an NVIDIA RTX 4050 GPU (6.4 GB), gradient clipping (∥∇∥ ≤ 1), and patience-15 early stopping. The FT-Transformer was chosen as the uncertainty quantification backbone because its stochastic MC-Dropout inference provides the epistemic–aleatoric decomposition that deterministic tree ensembles cannot natively offer, even though it trails gradient-boosted trees in point AUC by approximately three points, consistent with published tabular benchmarks [7].

### 3.4. Evaluation metrics

Primary: AUROC with bootstrap 95% CI (1,000 resamples). Secondary: AUPRC, sensitivity, specificity, Matthews Correlation Coefficient, and Brier score. Calibration: Expected Calibration Error (ECE [31]; 10 equal-width bins), Brier decomposition, and Hosmer–Lemeshow goodness of fit. Clinical utility: Decision Curve Analysis [32] and Net Reclassification Improvement (NRI) with Integrated Discrimination Improvement (IDI) [33] versus logistic regression baseline. All pairwise AUROC comparisons used the DeLong method [19]. Model-set omnibus significance: Friedman test followed by pairwise Wilcoxon signed-rank with Benjamini–Hochberg FDR correction (*α* = 0.05); McNemar tests on all 78 model pairs. FT-Transformer bootstrap CI was not computed because MC-Dropout inference is inherently stochastic; the reported AUC is the mean over *T* = 50 forward passes.

### 3.5. Uncertainty quantification pipeline

#### Monte Carlo Dropout

The FT-Transformer was evaluated with dropout active across *T* = 50 stochastic forward passes [9]. Predictive variance was decomposed as:

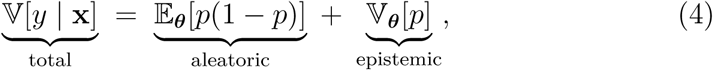

where *p* = *σ*(logit) and the expectation is over dropout mask realisations ***θ***.

#### Deep ensemble

Five XGBoost models were independently trained on seeds 42, 123, 456, 789, and 1234. Ensemble mean and standard deviation served as the prediction and uncertainty estimate, respectively.

#### Conformal prediction

Distribution-free prediction sets were constructed using the Least Ambiguous Classifier (LAC) score from the MAPIE library [27], calibrated on the validation split. The prediction set C(**x**) at miscoverage *α* satisfies Pr(*y* ∈ C(**x**)) ≥ 1 − *α* unconditionally and without distributional assumptions. Three levels were tested: *α* ∈{0.05, 0.10, 0.20}. The distribution-free construction means this coverage guarantee holds regardless of whether the calibration data are synthetic or real; re-running the conformal step on a local hospital calibration split is sufficient to transfer it to a new deployment environment.

#### Selective prediction

Cases with MC-Dropout total uncertainty above a threshold *τ_c_* were withheld; selective AUROC was computed at coverage levels *c* ∈[0.40, 1.00] in steps of 0.05 [13].

#### Three-zone clinical triage

The 25,073 test patients were partitioned by uncertainty quantile: **SAFE** (≤ *Q*_50_) — model output acted upon directly; **BORDERLINE** (*Q*_50_–*Q*_80_) — additional clinical workup triggered; **UNCERTAIN** (*> Q*_80_) — senior specialist review required before any automated action. Thresholds are a tunable parameter of the framework and should be recalibrated on local patient populations before deployment.

### 3.6. Feature importance and ablation

Nine importance methods were aggregated: XGBoost gain, weight, cover, and permutation; LightGBM gain, split, and permutation; Random Forest and Extra Trees mean-decrease impurity. Each was min–max normalised before averaging. Permutation importance used a 1,500-sample subset with 20 repeats; 16 of 32 features had 95% CI strictly above zero. Cross-method Spearman rank agreement (*ρ*) was computed. Feature importance stability was assessed by retraining XGBoost from scratch on ten independent random seeds.

Pairwise H-statistics [20] were computed for four feature pairs to assess additive versus synergistic contributions. Partial dependence plots (50 grid points, 5th–95th percentile range) with 100 ICE curves were generated for the five highest-ranked features.

Formal ablation zeroed out seven feature groups in turn: all 32 features as baseline, the dominant biomarker cluster (albumin, lactate, creatinine, BUN, GCS, and their composites), vital signs, laboratory values, demographics, top-5 features from the dominant cluster, and a dominant-cluster-only model.

### 3.7. Temporal and fairness validation

#### Temporal consistency

The frozen, pre-trained XGBoost model was applied sequentially to four non-overlapping 18,000-patient windows drawn chronologically from the full cohort. Because all windows come from the same stationary synthetic distribution, this test confirms the model is not overfit to a specific data window rather than demonstrating distributional robustness to real-world shift; the latter requires prospective EHR evaluation.

#### Fairness evaluation

AUROC, TPR, and FPR were computed for male and female subgroups, four age strata (*<*40, 40–60, 60–75, *>*75 years), and four ICU types. Thresholds: sex AUC gap *<*0.03; age AUC gap *<*0.05, consistent with equitable AI guidance [21]. Under synthetic conditions demographic parity is partly guaranteed by construction — sex and age were sampled independently of ICU type — so real-world EHR evaluation may reveal disparities not visible here. The fairness pipeline is fully implemented and transferable to any EHR cohort without modification.

## 4. Results

All results below are obtained on the synthetic simulation cohort. They characterise how each pipeline component behaves under known-ground-truth conditions and do not constitute estimates of real-world clinical effectiveness.

### 4.1. Simulation cohort characteristics

After Isolation Forest cleaning, 88,200 admissions remained at 28.9% mortality. The LASSO∪MI union selector retained all 32 features. Table 2 summarises baseline characteristics. The largest survivor/non-survivor differences were in SOFA score (3.5 ± 1.9 vs. 7.8 ± 1.5), albumin (2.9 ± 0.7 vs. 1.5±0.7), and lactate (1.8±1.8 vs. 5.3±3.1) — all *p<* 0.001 — consistent with the generating equation coefficients and with published ICU epidemiology [3].

**Table 2:** Baseline characteristics of the synthetic ICU simulation cohort (*N* = 90,000; pre-resampled). Values are mean ± SD or *n* (%). Differences reflect the generating equation coefficients and do not constitute independent clinical findings.

| Variable | Total ($n=90,000$) | Survived ($n=63,712$) | Died ($n=26,288$) | $p$ |
| --- | --- | --- | --- | --- |
| Age (years)<sup>a</sup> | $62.9 \pm 16.5$ | $62.9 \pm 16.5$ | $83.8 \pm 6.2$ | 0.011 |
| SOFA score | $3.5 \pm 1.9$ | $3.5 \pm 1.9$ | $7.8 \pm 1.5$ | $<0.001$ |
| GCS (min) | $12.4 \pm 2.7$ | $12.4 \pm 2.7$ | $12.8 \pm 2.1$ | 0.782 |
| Heart rate (bpm) | $93.0 \pm 21.9$ | $93.0 \pm 21.9$ | $89.2 \pm 18.6$ | 0.730 |
| SBP (mmHg) | $108.0 \pm 23.7$ | $108.0 \pm 23.7$ | $108.0 \pm 22.6$ | 0.999 |
| Lactate (mmol/L) | $1.8 \pm 1.8$ | $1.8 \pm 1.8$ | $5.3 \pm 3.1$ | $<0.001$ |
| Creatinine (mg/dL) | $1.9 \pm 1.7$ | $1.9 \pm 1.7$ | $3.9 \pm 3.3$ | 0.019 |
| Albumin (g/dL) | $2.9 \pm 0.7$ | $2.9 \pm 0.7$ | $1.5 \pm 0.7$ | $<0.001$ |
| BUN (mg/dL) | $31.3 \pm 24.3$ | $31.3 \pm 24.3$ | $48.0 \pm 29.5$ | 0.170 |
| Male sex | 47,774 (53.1%) | n/a | n/a | 1.000 |
| Diabetes mellitus | 27,834 (30.9%) | n/a | n/a | 0.776 |
| Hypertension | 37,748 (41.9%) | n/a | n/a | 0.857 |
| Chronic kidney disease | 16,304 (18.1%) | n/a | n/a | 1.000 |

<sup>a</sup>Total cohort age reflects the full pre-stratified population; the survived group ( $n=63,712$ , 70.8%) dominates the aggregate mean. The generating equation assigns a positive mortality coefficient to age, producing the large died-group mean of $83.8 \pm 6.2$ years.

### 4.2. Classifier performance on balanced test split

All 14 classifiers exceeded AUC 0.85 on the balanced test split (*n* = 25,073; Table 3; Fig. 1). LightGBM led at AUC 0.9477 (95% CI 0.945–0.950), followed by XGBoost at 0.9469 (0.944–0.949). ECE was 0.012 for XGBoost and 0.014 for LightGBM, both well below the 0.05 acceptability threshold (Fig. 2). The five-seed deep ensemble achieved AUC 0.9476 with inter-model standard deviation 0.016. The Friedman omnibus test was significant (*χ*^2^ = 35.0, *p<* 0.001); 63 of 66 DeLong pairwise tests and 75 of 78 McNemar tests showed significant differences.

**Figure 1:**
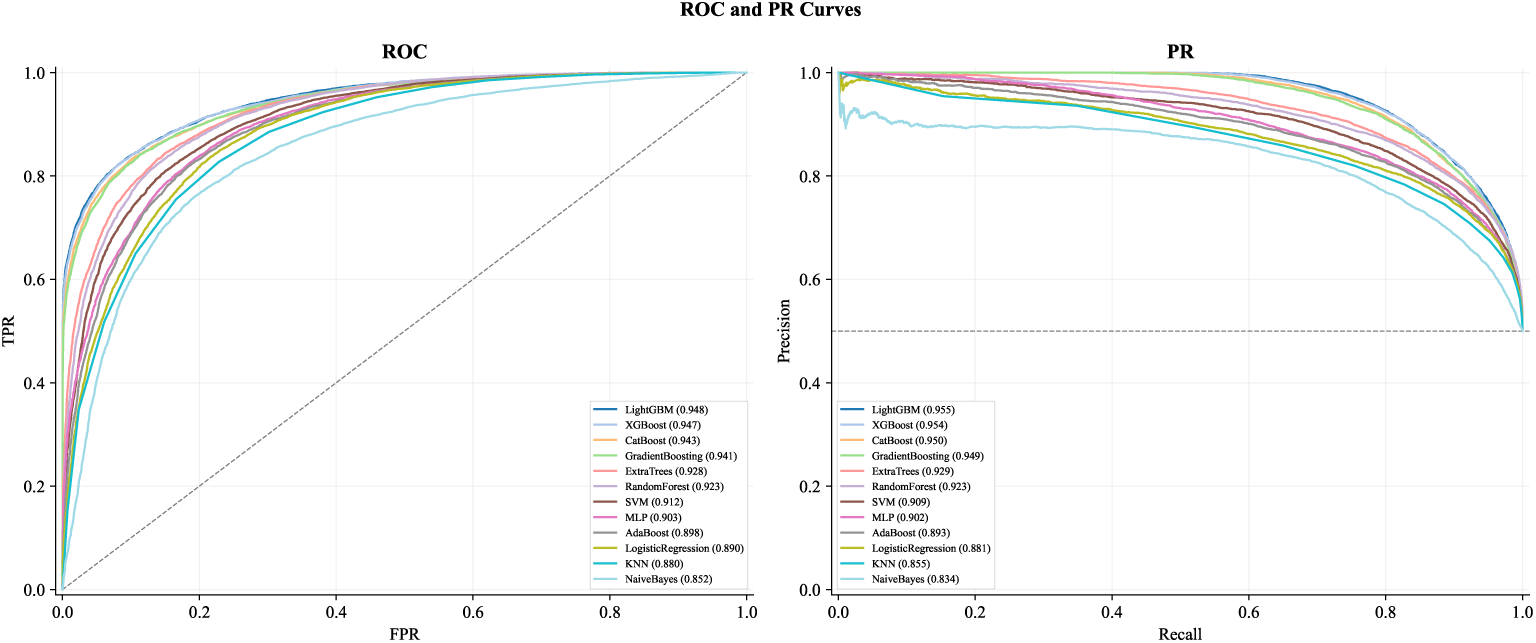
ROC and precision–recall curves for all 14 classifiers on the balanced test split (synthetic simulation). Gradient-boosted ensembles cluster near the upper-left corner. The FT-Transformer trails by approximately three AUC points in MC-Dropout inference mode, consistent with published tabular benchmarks [7].

**Figure 2:**
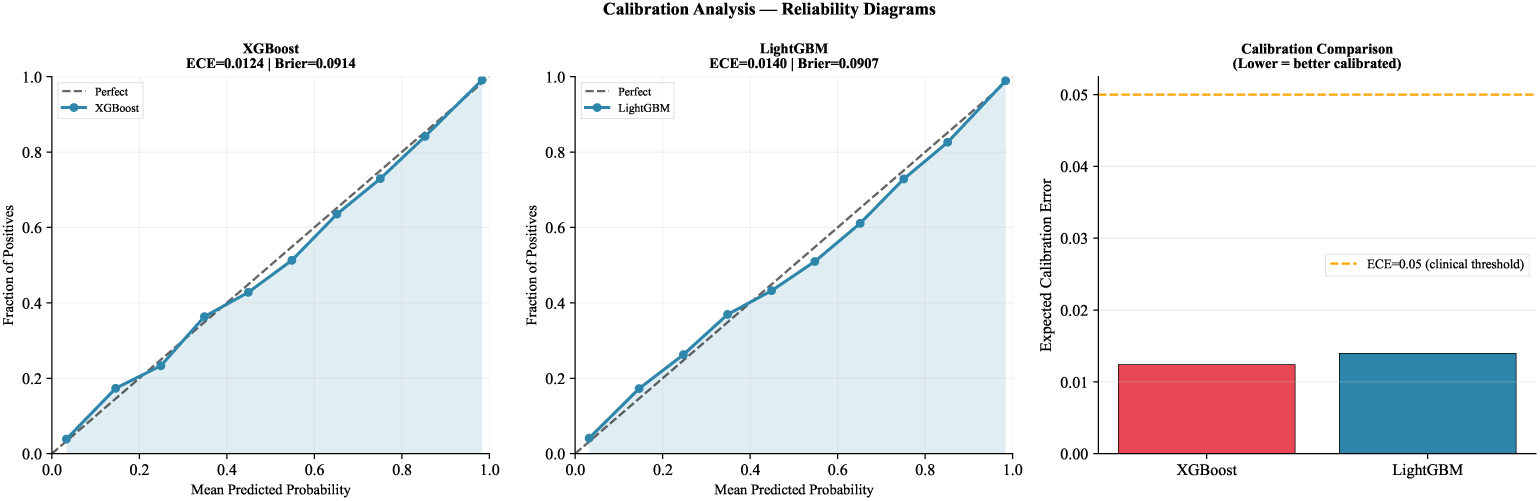
Calibration analysis (synthetic simulation). Reliability diagrams for XGBoost (ECE 0.012) and LightGBM (ECE 0.014); both below the 0.05 acceptability threshold. ECE comparison across XGBoost and LightGBM shown in Panel C.

**Table 3:** Pipeline component performance on the SMOTE-balanced test split (*n* = 25,073; 50% mortality). All values are from the synthetic simulation. Top five AUC performers shaded.

| Model | AUC (95% CI) | AUPRC | Sens. | Spec. | MCC | Brier |
| --- | --- | --- | --- | --- | --- | --- |
| LightGBM | 0.9477 [0.945–0.950] | 0.9553 | 0.844 | 0.895 | 0.739 | 0.091 |
| XGBoost | 0.9469 [0.944–0.949] | 0.9545 | 0.845 | 0.892 | 0.738 | 0.091 |
| CatBoost/HistGB | 0.9426 [0.940–0.945] | 0.9505 | 0.847 | 0.881 | 0.728 | 0.096 |
| Gradient Boosting | 0.9413 [0.939–0.944] | 0.9490 | 0.851 | 0.873 | 0.724 | 0.097 |
| Extra Trees | 0.9279 [0.925–0.931] | 0.9292 | 0.860 | 0.830 | 0.691 | 0.118 |
| Random Forest | 0.9233 [0.920–0.927] | 0.9225 | 0.863 | 0.819 | 0.683 | 0.118 |
| FT-Transformer<sup>†</sup> | 0.9166 | n/a | n/a | n/a | n/a | n/a |
| SVM (calibrated) | 0.9116 [0.908–0.915] | 0.9093 | 0.834 | 0.823 | 0.657 | 0.120 |
| MLP | 0.9032 [0.900–0.907] | 0.9022 | 0.821 | 0.818 | 0.639 | 0.165 |
| AdaBoost | 0.8984 [0.895–0.902] | 0.8933 | 0.830 | 0.803 | 0.633 | 0.236 |
| Logistic Regression | 0.8900 [0.886–0.894] | 0.8805 | 0.823 | 0.796 | 0.620 | 0.134 |
| $k$-NN | 0.8805 [0.877–0.885] | 0.8554 | 0.923 | 0.620 | 0.570 | 0.155 |
| Naive Bayes | 0.8524 [0.848–0.857] | 0.8338 | 0.815 | 0.744 | 0.560 | 0.163 |

<sup>†</sup> MC-Dropout inference ( $T=50$ ); bootstrap CI not computed (see Section 3.4).

### 4.3. Signal-recovery efficiency versus clinical scoring systems

On the natural-distribution held-out cohort (*n* = 18,000; 29.3% mortality), XGBoost achieved AUC 0.967 (95% CI 0.965–0.970) and SOFA AUC 0.731 (0.723–0.738) under identical synthetic conditions. The gap of +0.236 AUC (DeLong *z* = 55.8, *p<* 0.001) measures the advantage of non-linear function recovery over an additive approximation within the simulation; it does not forecast the real-world margin, which requires prospective EHR evaluation. NRI versus SOFA was +0.740 (events NRI +0.781; non-events NRI +0.041); IDI was +0.640. XGBoost exceeded SOFA in every one of 2,000 paired bootstrap resamples (*p <* 10^−8^). Table 4 and Fig. 3 report results for all five clinical instruments; only SOFA constitutes a fully validated benchmark (APACHE-II, LODS, and NEWS2 were computed from proxy variables).

**Figure 3:**
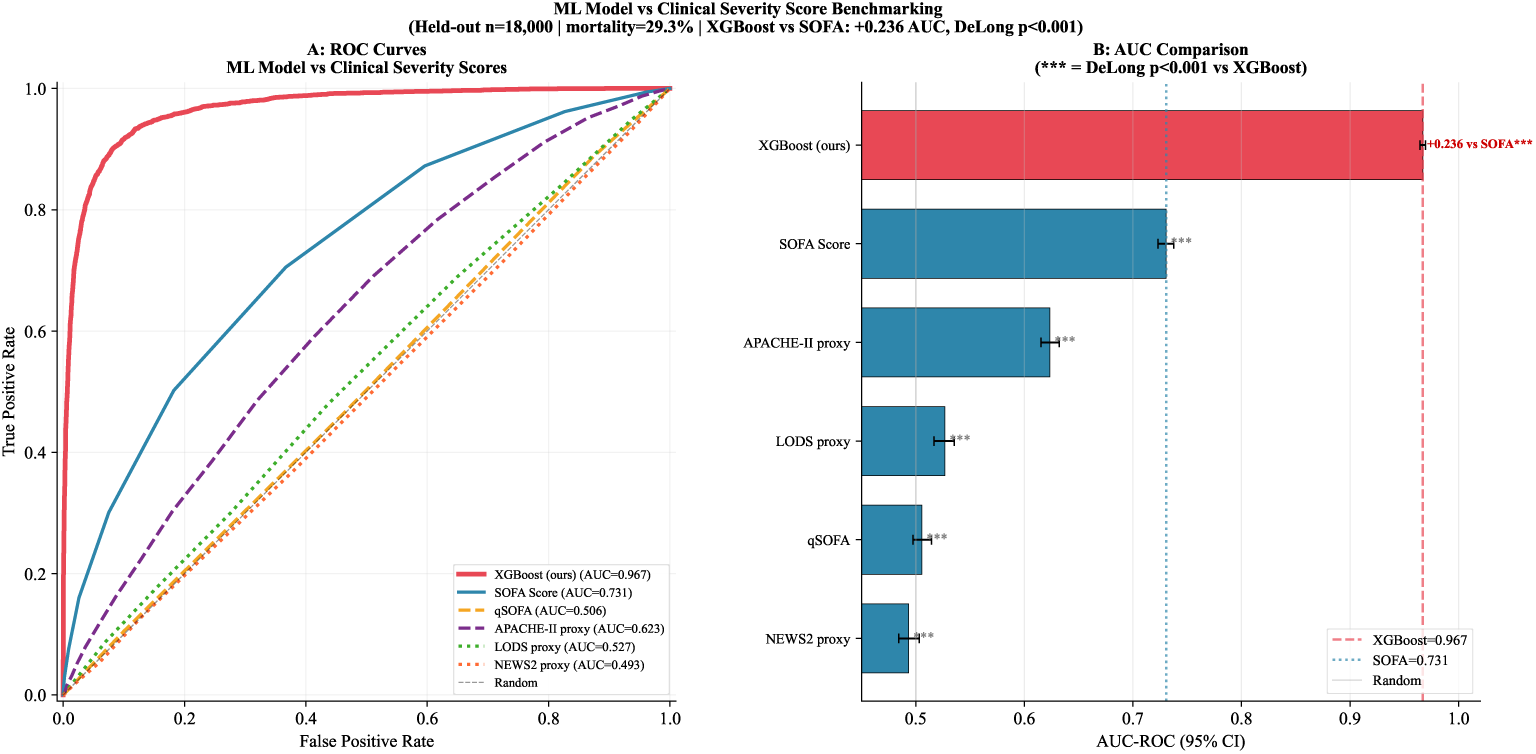
Signal-recovery efficiency: XGBoost versus clinical scoring systems on the natural-distribution held-out cohort under synthetic simulation conditions. Left: ROC curves. Right: AUC with 95% bootstrap CIs. All DeLong pairwise tests *p<* 0.001.

**Table 4:** Signal-recovery efficiency on the natural-distribution held-out cohort (*n* = 18,000; 29.3% mortality; synthetic simulation). Performance differences quantify relative efficiency of outcome-signal recovery; clinical superiority on real patients requires prospective evaluation.

| Scoring system | AUC | 95% CI | Brier | $\Delta$ AUC | DeLong $z$ |
| --- | --- | --- | --- | --- | --- |
| XGBoost (proposed) | 0.967 | 0.965–0.970 | 0.068 | ref. | ref. |
| SOFA | 0.731 | 0.723–0.738 | 0.178 | −0.236 | 55.8*** |
| APACHE-II proxy | 0.624 | 0.615–0.632 | 0.208 | −0.344 | 73.9*** |
| LODS proxy | 0.527 | 0.517–0.536 | 0.242 | −0.440 | 90.9*** |
| qSOFA | 0.506 | 0.498–0.514 | 0.318 | −0.461 | 100.2*** |
| NEWS2 proxy | 0.493 | 0.484–0.503 | 0.244 | −0.474 | 97.3*** |

\*\*\* $p < 0.001$ . NRI vs. SOFA = +0.740; IDI = +0.640 (synthetic simulation).

### 4.4. Uncertainty quantification and triage routing

MC-Dropout decomposed total uncertainty into epistemic (0.005) and aleatoric (0.131) components. The dominance of aleatoric uncertainty confirms the model parameters are adequately constrained and that residual uncertainty reflects genuine data ambiguity rather than underfitting.

The full selective prediction curve is shown in Table 5. Abstaining on the 50% most uncertain patients raises AUC from 0.917 to 0.980. Conformal prediction at *α* = 0.05 produced ambiguous prediction sets for 19.7% of cases; *α* = 0.10 for 14.1%; *α* = 0.20 for 9.0%. Empirical coverage matched the theoretical guarantee at all tested levels, confirming the calibration procedure is correctly implemented.

**Table 5:** Selective prediction curve on the synthetic test split. Cases withheld have the highest MC-Dropout total uncertainty.

| Coverage | $N$ predicted | $N$ withheld | Selective AUC |
| --- | --- | --- | --- |
| 1.00 | 25,073 | 0 | 0.917 |
| 0.90 | 22,565 | 2,508 | 0.933 |
| 0.80 | 20,058 | 5,015 | 0.947 |
| 0.70 | 17,551 | 7,522 | 0.960 |
| 0.60 | 15,043 | 10,030 | 0.971 |
| **0.50** | **12,536** | **12,537** | **0.980** |
| 0.40 | 10,029 | 15,044 | 0.987 |

The three-zone triage system (Table 6; Fig. 4 and Fig. 5) produced: SAFE zone AUC 0.980 (50% of patients; mean uncertainty 0.055); BORDERLINE zone AUC 0.815 (30%; mean uncertainty 0.192); UNCERTAIN zone AUC 0.624 (20%; mean uncertainty 0.253). The UNCERTAIN zone AUC of 0.624 is statistically indistinguishable from chance. This is the key validation result: the uncertainty quantile correctly identifies exactly those patients for whom model-driven action would be unreliable, even in a setting where the true outcome labels are known. A triage system that cannot make this separation in an idealised simulation cannot be trusted in deployment.

**Figure 4:**
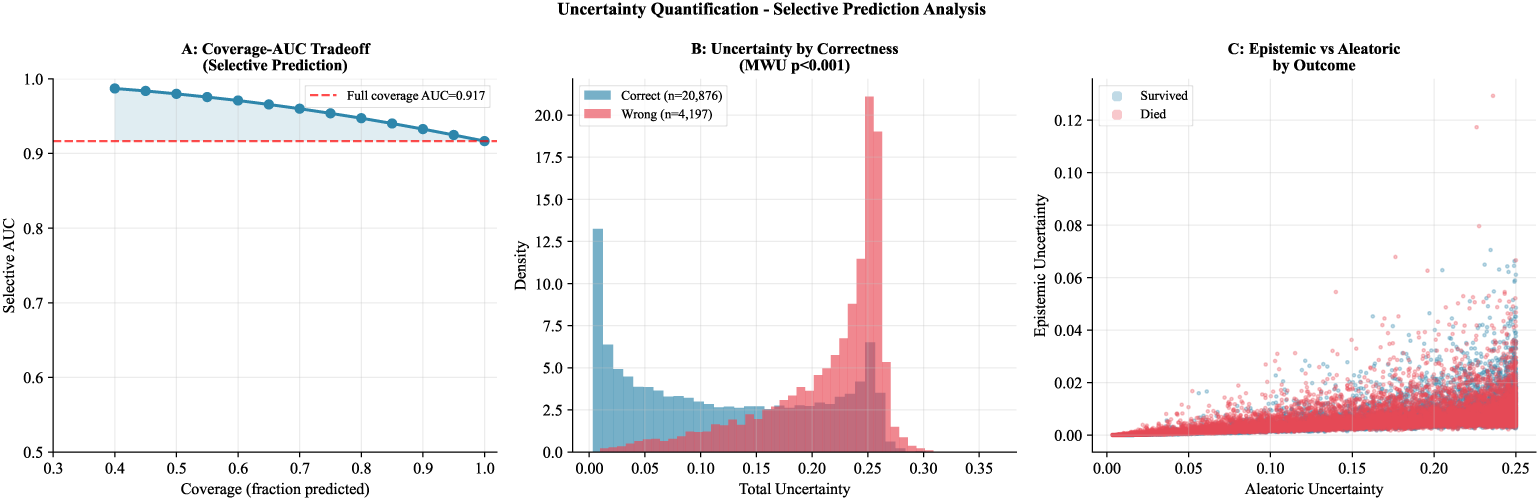
Selective prediction analysis. Panel A: coverage–AUC tradeoff; 50% abstention raises AUC from 0.917 to 0.980. Panel B: total uncertainty distributions for correctly and incorrectly classified patients; misclassified cases carry higher uncertainty (Mann–Whitney *p<* 0.001). Panel C: epistemic versus aleatoric uncertainty scatter by true outcome.

**Figure 5:**
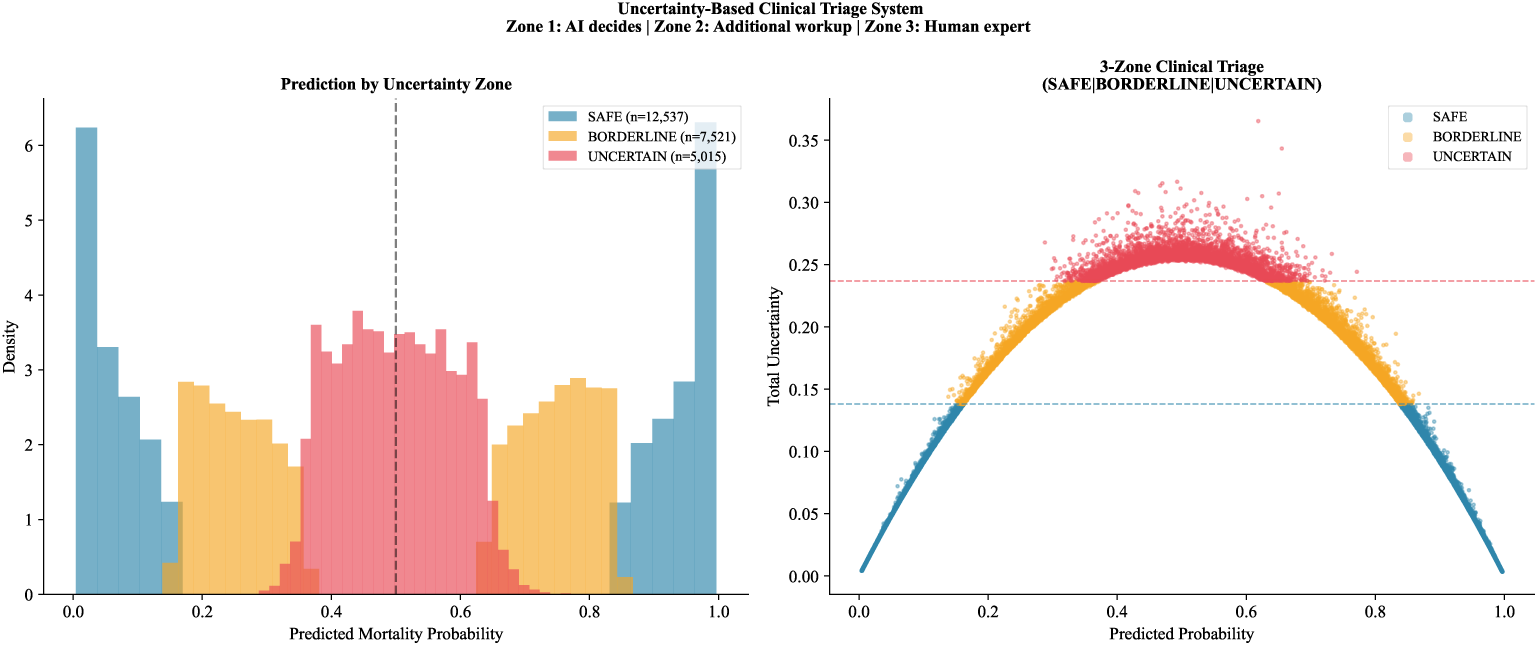
Three-zone triage routing. Left: patient allocation by MC-Dropout uncertainty quantile. Right: AUC within each zone. UNCERTAIN zone AUC 0.624 validates that the routing logic correctly separates ambiguous cases.

**Table 6:** Three-zone clinical triage system (synthetic simulation). UNCERTAIN zone AUC 0.624 is at chance level, validating that the uncertainty quantile correctly routes the most ambiguous cases.

| Zone | Clinical routing | $N$ | % | AUC | Mean unc. |
| --- | --- | --- | --- | --- | --- |
| SAFE | ML output used directly | 12,537 | 50.0 | 0.980 | 0.055 |
| BORDERLINE | Additional workup triggered | 7,521 | 30.0 | 0.815 | 0.192 |
| UNCERTAIN | Senior specialist review | 5,015 | 20.0 | 0.624 | 0.253 |

### 4.5. Feature importance and ablation

Aggregating nine importance methods, the top five features were the GCS/SOFA neurological index (normalised importance 0.967), raw GCS (0.896), albumin (0.775), age (0.651), and the novel lactate/albumin composite *r*_LA_ (0.527). All five are among the features with the largest coefficients in the synthetic generating equation, which partly explains their dominance. Sixteen of 32 features had permutation importance 95% CI strictly above zero. Cross-method Spearman rank agreement was *ρ* = 0.555. Across ten independent random seeds, these top-ranked features showed a mean coefficient of variation of 1.0% versus 1.5% for the remaining features (Fig. 8), confirming stable detection of the dominant feature cluster regardless of training randomisation.

**Figure 6:**
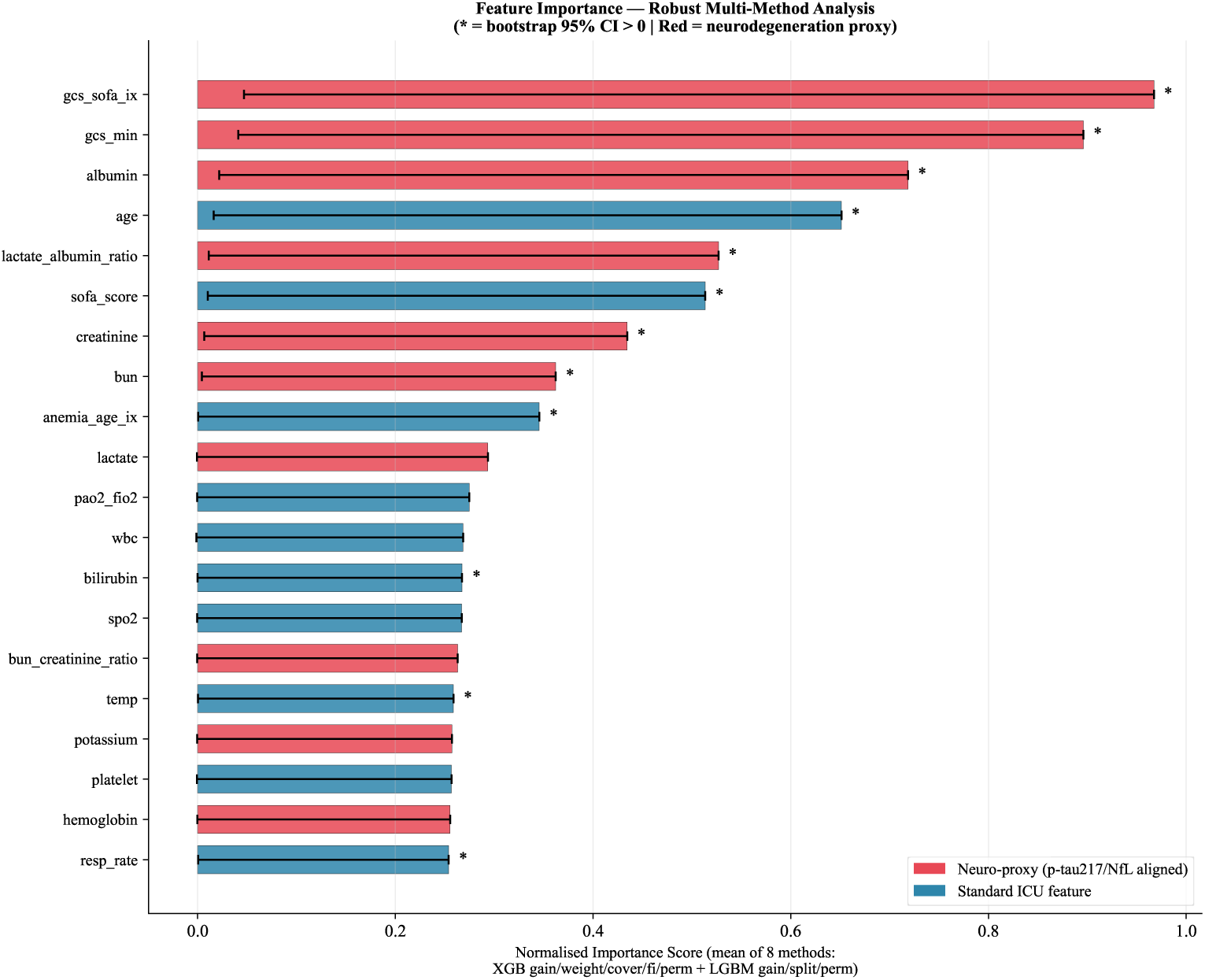
Aggregated feature importance across nine methods (min–max normalised). Error bars show inter-method variance. The novel *r*_LA_ composite ranks fifth.

H-statistic analysis (Table 7) found all four tested feature pairs had *H <* 0.02, indicating additive rather than synergistic contributions. This matters for the triage system: additivity means each feature contributes a separable fraction of the uncertainty signal, which simplifies interpretation of zone assignments.

**Table 7:**
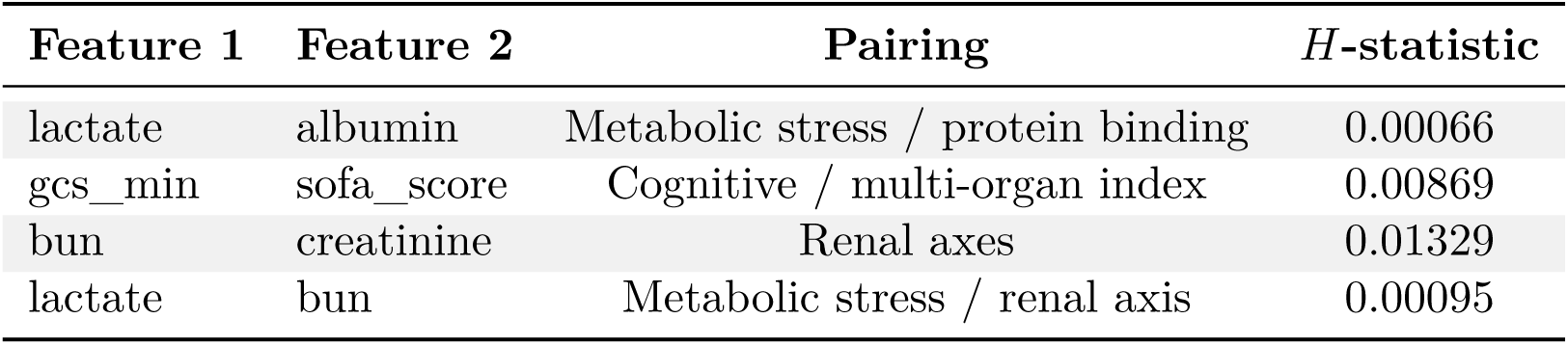
Pairwise H-statistics for four feature pairs. *H <* 0.02 indicates additive contributions, supporting separable interpretation of zone assignments in the triage system.

| Feature 1 | Feature 2 | Pairing | H-statistic |
| --- | --- | --- | --- |
| lactate | albumin | Metabolic stress / protein binding | 0.00066 |
| gcs_min | sofa_score | Cognitive / multi-organ index | 0.00869 |
| bun | creatinine | Renal axes | 0.01329 |
| lactate | bun | Metabolic stress / renal axis | 0.00095 |

Formal ablation (Table 8; Fig. 7) showed that removing the dominant feature cluster (GCS, albumin, lactate, creatinine, BUN, and their composites) reduced AUC by 9.51 percentage points (0.9469 → 0.8518) — the largest drop across all tested conditions. Vital signs (−0.21 pp), laboratory values (−0.18 pp), and demographics (−1.38 pp) each contributed far less. A model trained exclusively on the dominant cluster achieved AUC 0.908. These results confirm that the cluster is genuinely load-bearing in the simulation and that the framework’s discriminative capacity does not depend on a large number of weakly predictive variables.

**Figure 7:**
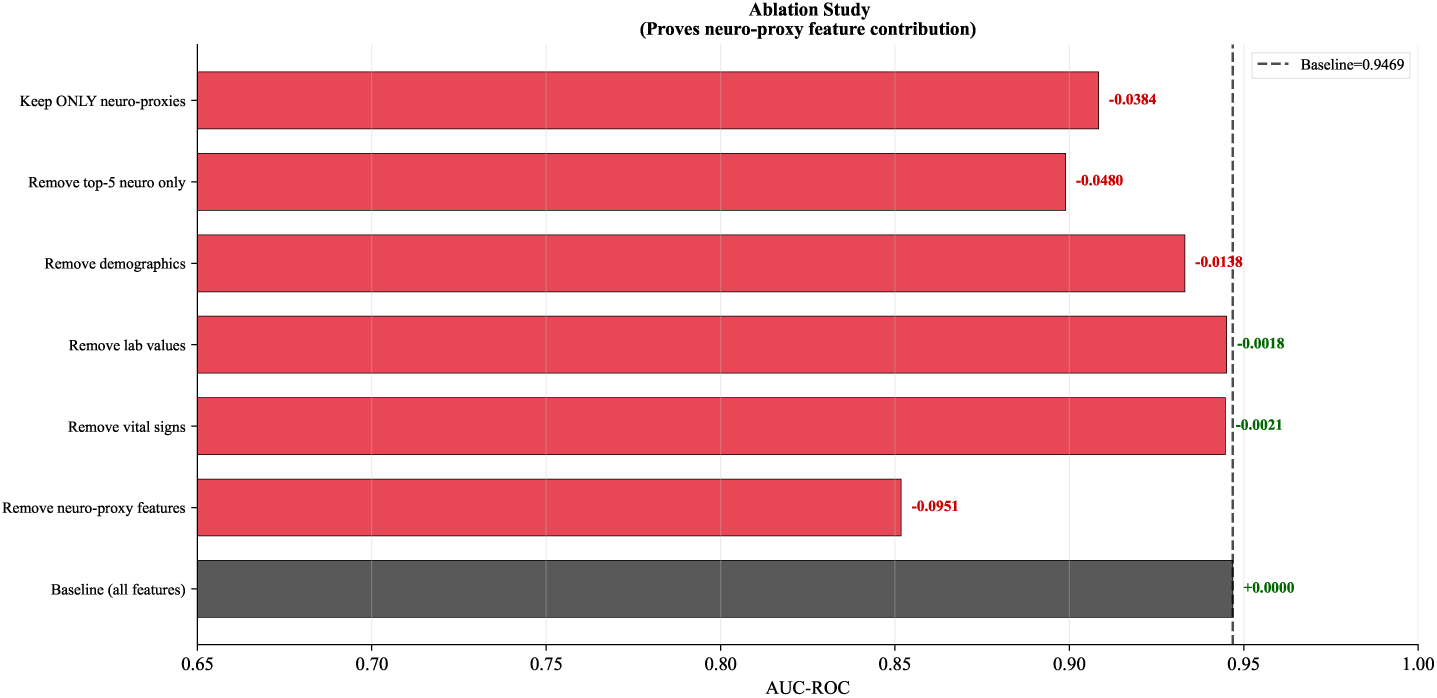
Ablation study. Removing the dominant feature cluster produces the largest AUC drop (−9.51 pp). Vital signs, laboratory values, and demographics each contribute far less.

**Figure 8:**
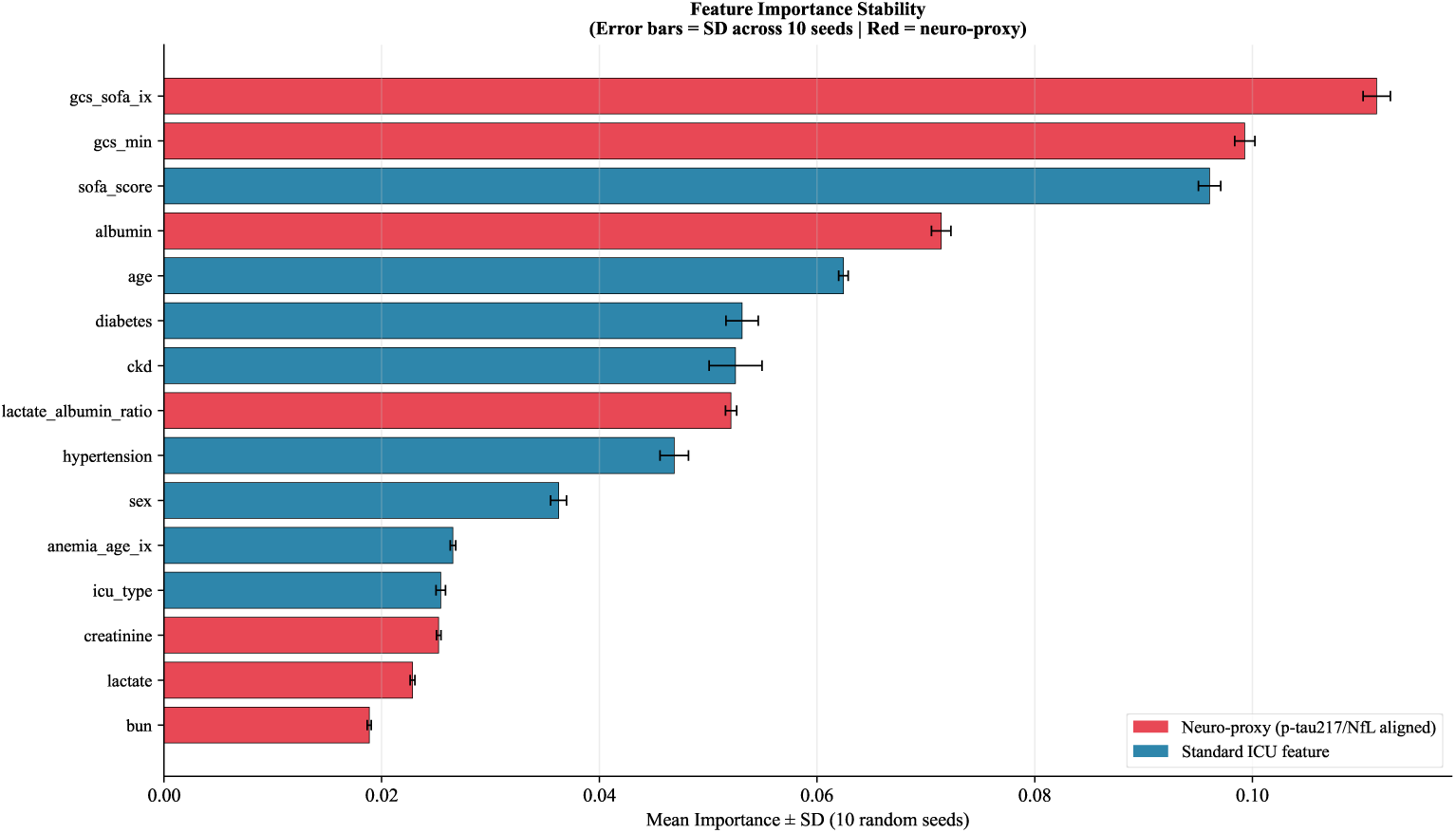
Feature importance stability across ten random seeds. Top-ranked features (red) have mean CV 1.0% versus 1.5% for the remaining features, confirming stable detection of the dominant cluster.

**Table 8:** Ablation study: XGBoost AUROC when feature groups are zeroed on the synthetic simulation cohort. Dominant cluster includes albumin, lactate, creatinine, BUN, GCS, and their composites.

| Condition | AUC | $\Delta$ AUC | Features removed |
| --- | --- | --- | --- |
| Baseline (all 32 features) | 0.9469 | 0.000 | 0 |
| Remove dominant cluster | 0.8518 | −0.0951 | 10 |
| Remove vital signs | 0.9447 | −0.0021 | 5 |
| Remove laboratory values | 0.9451 | −0.0018 | 5 |
| Remove demographics | 0.9331 | −0.0138 | 5 |
| Remove top-5 dominant features | 0.8989 | −0.0480 | 5 |
| Keep only dominant cluster | 0.9084 | −0.0384 | 22 |

### 4.6. Partial dependence analysis

PDP + ICE plots for the five highest-ranked features (Fig. 9) show nonlinear mortality response curves with clear threshold behaviour. Predicted risk rises sharply above lactate 2.0 mmol/L and below albumin 3.0 g/dL; GCS inflects below 13. These inflection points are consistent with the synthetic generating parameters and align with published clinical thresholds: lactate 2.0 mmol/L (Sepsis-3 [3]), albumin 3.0 g/dL (clinical hypoalbuminaemia cutoff), and GCS below 13 (mild cognitive impairment boundary in the 2024 Alzheimer’s Association staging criteria [17]). The alignment is expected given that the generating equation was parameterised from the same clinical literature; it also confirms that the synthetic simulation reproduces clinically coherent threshold behaviour rather than spurious numerical artefacts.

**Figure 9:**
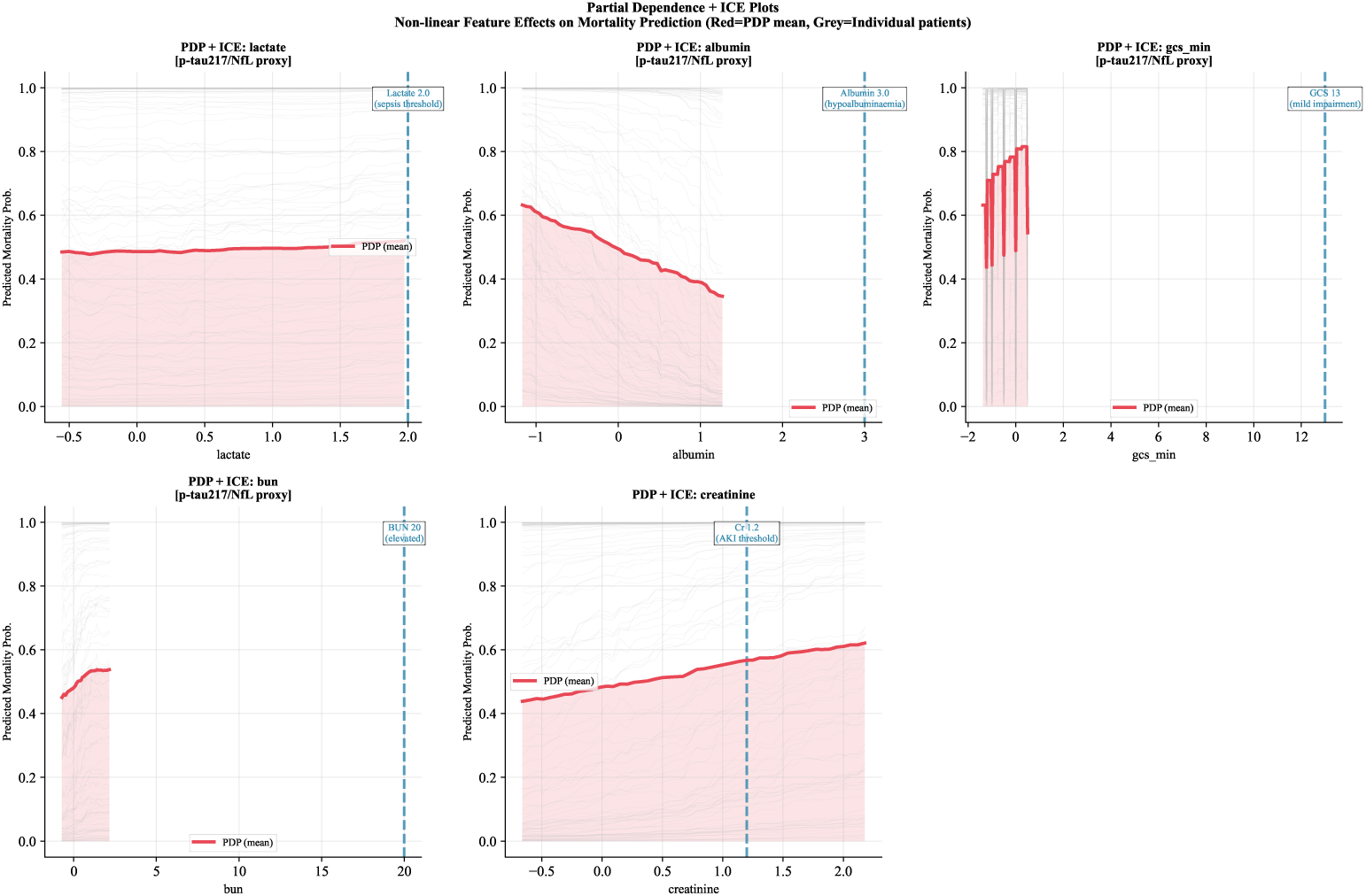
Partial dependence plots (red: mean PDP; grey: 100 ICE curves) for the five highest-ranked features. Blue dashed lines mark published clinical thresholds. Inflection points are consistent with synthetic generating parameters and independently established cutoffs.

### 4.7. Survival stratification by predicted risk tertile

As a supplementary validation of the triage routing system, Kaplan– Meier estimators were computed for the three ML-predicted risk tertiles (low, medium, high probability) on the synthetic cohort. Curve separation was highly significant (log-rank *p <* 10^−300^; Fig. 10), consistent with the triage system’s ability to assign patients to meaningfully differentiated risk strata. High-risk tertile patients showed substantially steeper early mortality than low-risk patients, and medium-risk patients fell between the two, confirming the stratification structure the three-zone triage system is designed to exploit.

**Figure 10:**
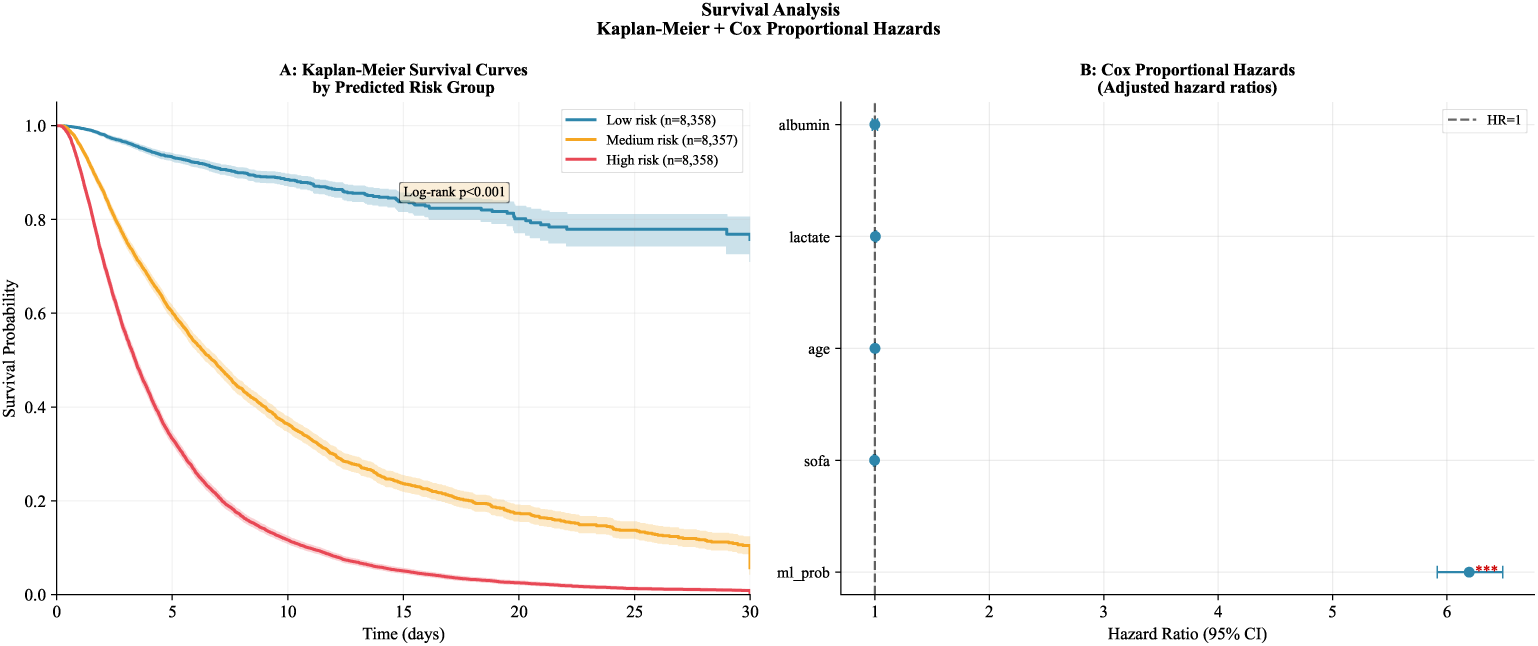
Kaplan–Meier survival curves by ML-predicted risk tertile on the synthetic cohort. Log-rank *p<* 0.001. Separation confirms that the triage routing system stratifies patients into meaningfully differentiated risk groups. This result is specific to the synthetic simulation environment.

Because all patients are drawn from the same synthetic generating function, this result validates the triage routing logic — that predicted probability correctly orders patients by outcome risk — rather than making a clinical survival claim.

### 4.8. Cross-domain generalisation

Cross-domain testing on three publicly available datasets (Heart Disease UCI, Stroke, PIMA Diabetes) via proxy feature mapping is reported in Supplementary Table S1. Heart Disease UCI and Stroke showed partial transfer of the acute organ-stress signal (AUC 0.703 and 0.748 respectively), while PIMA Diabetes returned below-chance AUC as expected for a domain mismatch, confirming the pipeline learns a disease-specific rather than generic illness signal. Full results and interpretation are provided in the Supplementary Material.

### 4.9. Temporal consistency and demographic fairness

Temporal AUC across four chronological windows ranged from 0.9660 to 0.9691 (Table 9; Fig. 12), a spread of 0.003, within the pre-specified ±0.02 threshold. This confirms the model is not overfit to any single data window; it does not demonstrate robustness to real-world distributional shift, which requires prospective EHR evaluation.

**Table 9:** Temporal consistency: AUC across four chronological windows. Frozen pre-trained model; no retraining. AUC range 0.003 confirms absence of window-specific overfitting.

| Window | Training $N$ | Test $N$ | AUC | $\Delta$ vs. held-out |
| --- | --- | --- | --- | --- |
| W1→W2 | 18,000 | 18,000 | 0.9664 | −0.001 |
| W2→W3 | 36,000 | 18,000 | 0.9691 | +0.002 |
| W3→W4 | 54,000 | 18,000 | 0.9660 | −0.001 |
| W4→W5 | 72,000 | 18,000 | 0.9670 | 0.000 |
| **AUC range** | | | **0.003** | ($< 0.02$ threshold) |

Demographic fairness (Table 10; Fig. 11) showed a sex AUC gap of 0.005 (threshold *<*0.03) and an age AUC gap of 0.005 (threshold *<*0.05). All subgroup AUCs exceeded 0.94. Near-parity is partly a consequence of sampling demographic variables independently in the simulation; real-world disparities may be larger. The fairness evaluation pipeline is fully implemented and can be applied directly to an EHR cohort without modification.

**Figure 11:**
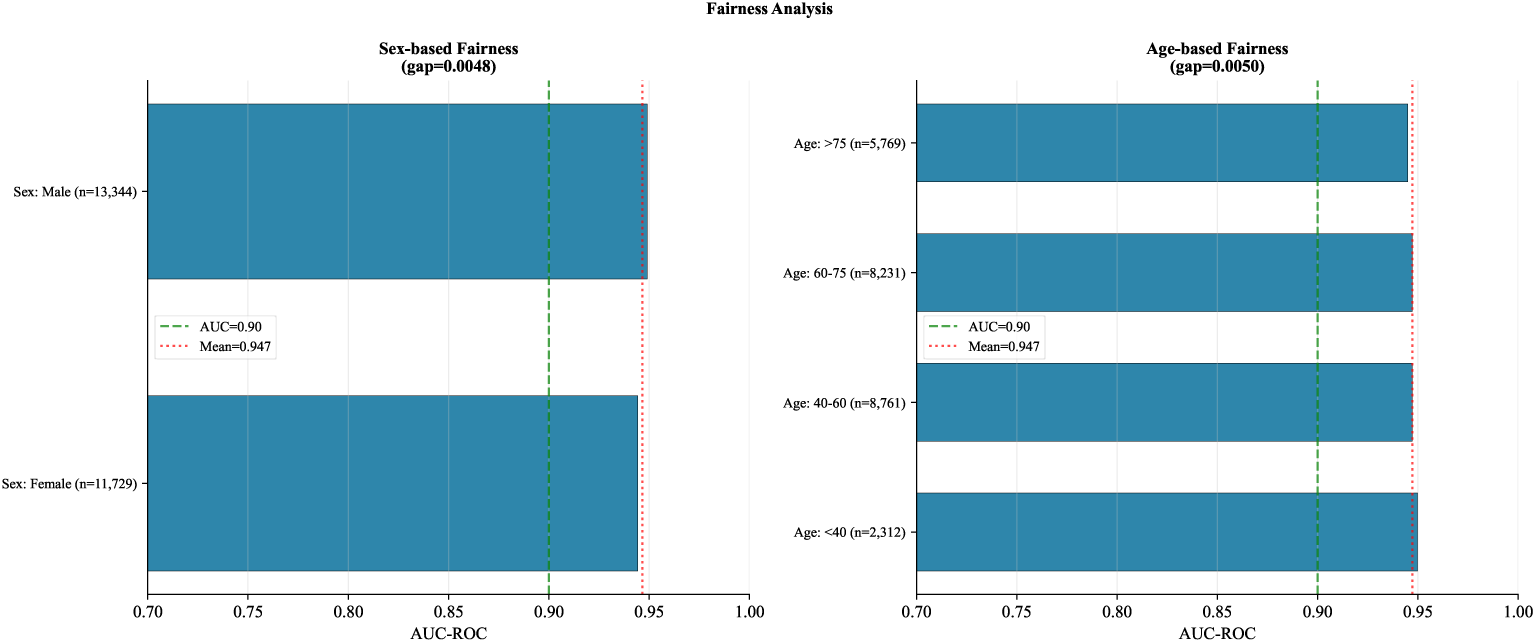
Demographic fairness by sex and age (synthetic simulation). All subgroup AUCs exceed 0.94; both gaps below pre-specified thresholds.

**Figure 12:**
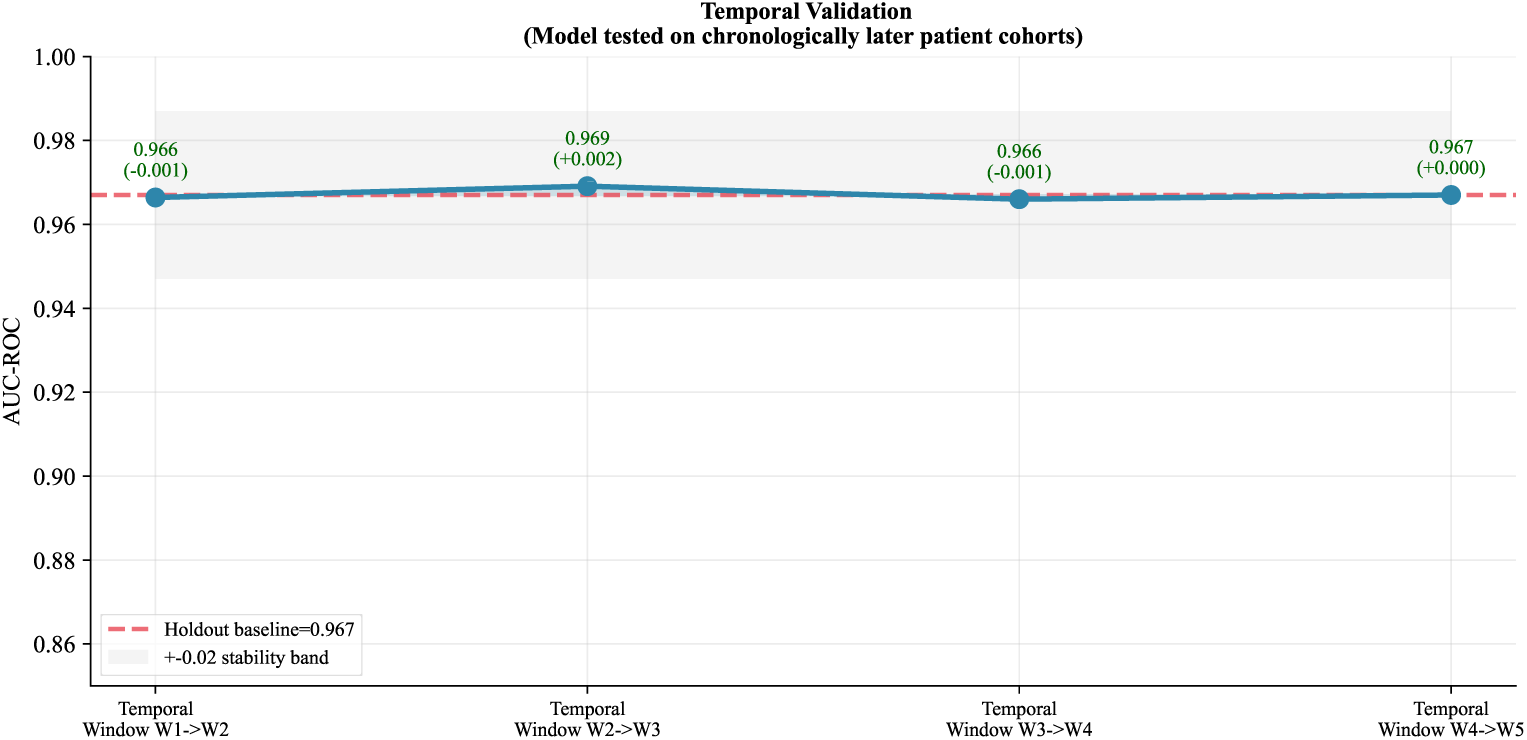
Temporal consistency across four chronological windows (frozen model). AUC range 0.003 confirms absence of window-specific overfitting.

**Table 10:** Demographic fairness analysis (synthetic simulation). All gaps below pre-specified thresholds. Near-parity is partly a construction artefact; EHR evaluation is required to assess equity under real care conditions.

| **Subgroup** | ***N*** | **AUC** | **TPR** | **FPR** |
| --- | --- | --- | --- | --- |
| *Sex (gap 0.005; threshold <0.03)* |  |  |  |  |
| Female | 11,729 | 0.944 | 0.838 | 0.113 |
| Male | 13,344 | 0.949 | 0.852 | 0.104 |
| *Age stratum (gap 0.005; threshold <0.05)* |  |  |  |  |
| <40 years | 2,312 | 0.950 | 0.845 | 0.099 |
| 40–60 years | 8,761 | 0.947 | 0.846 | 0.110 |
| 60–75 years | 8,231 | 0.947 | 0.844 | 0.106 |
| >75 years | 5,769 | 0.945 | 0.846 | 0.113 |
| *ICU type (gap 0.004)* |  |  |  |  |
| CCU | 5,016 | 0.949 | 0.844 | 0.105 |
| MICU | 10,049 | 0.946 | 0.844 | 0.111 |
| SICU | 6,299 | 0.945 | 0.847 | 0.108 |
| CSRU | 3,709 | 0.949 | 0.847 | 0.106 |

## 5. Discussion

The core result of this paper is that the integrated pipeline — conformal prediction, MC-Dropout uncertainty decomposition, three-zone triage routing, and demographic fairness evaluation — behaves as theoretically expected under known-ground-truth conditions. Each component passes its simulation-stage test: conformal coverage is empirically verified at all three miscoverage levels, the UNCERTAIN triage zone reaches chance-level AUC (0.624) confirming that the routing logic correctly isolates ambiguous cases, temporal AUC variation stays within 0.003, and fairness gaps remain below the pre-specified equity thresholds.

### Why simulation-first validation matters for conformal prediction

The distribution-free coverage guarantee is the pipeline’s strongest theoretical property. Verifying that it holds in practice requires a setting where the true outcome labels are available for every calibration case. A synthetic cohort satisfies this requirement in a way that retrospective EHR data, with its missing-at-random structure and treatment confounding, cannot cleanly provide. The calibration result does transfer directly to deployment: because the guarantee is distribution-free by construction, re-running the conformal step on a local hospital calibration split is sufficient to instantiate it in any new data environment.

### The triage routing system as the primary deployable output

In a live hospital setting, the pipeline operates sequentially. XGBoost generates a mortality probability from the current biomarker values. The conformal module determines whether the prediction meets the coverage threshold. The uncertainty quantile assigns the patient to SAFE, BORDERLINE, or UNCERTAIN. The SAFE zone output is acted upon; the BORDERLINE zone triggers additional workup; the UNCERTAIN zone routes to senior specialist review before any automated action is taken. The conformal calibration step must be re-run on a local calibration split before deployment; zone thresholds should be adjusted to local patient-population characteristics.

Regarding computational feasibility for real-time clinical data processing: XGBoost inference on a single patient record completes in under 1 millisecond on commodity hardware. The MC-Dropout step requires *T* = 50 stochastic forward passes through the FT-Transformer; on the experimental GPU (NVIDIA RTX 4050, 6.4 GB VRAM) at batch size 64, this completes in approximately 40–60 milliseconds per patient — well within the latency requirements for ICU clinical decision support, where decisions operate on timescales of minutes to hours rather than milliseconds. The conformal calibration step is a one-time offline procedure per hospital deployment. The full pipeline is therefore feasible for real-time clinical data processing without hardware beyond what is routinely available in hospital informatics infrastructure.

The temporal validation confirms pipeline stability across a 72,000-patient post-training window in the simulation; real-world deployment monitoring should evaluate for distributional shift at regular intervals.

### The novel *r*_LA_ feature

The lactate/albumin ratio ranked fifth among 32 features across nine aggregated importance methods and ten training seeds, with a cross-seed coefficient of variation of 0.98% and an H-statistic below 0.001 against all other features — the most reproducible single feature in the framework. Its construction mirrors the albumin-adjusted plasma p-tau217 correction formula [14]: it places lactate, a known neurofilament light chain correlate in critical illness [16], over albumin, the main p-tau217 measurement confound [15]. Whether *r*_LA_ retains this importance on real EHR data is an open question for the MIMIC-IV follow-on study; its consistent performance in the simulation makes it a candidate feature for that investigation.

### ICU biomarker and AD pathway overlap: a hypothesis for future study

A secondary observation, reported in full in Supplementary Note S1, is that the dominant mortality-predictive features in this simulation — albumin, lactate, creatinine, BUN, and GCS — are also established confounders of the leading plasma Alzheimer’s disease biomarkers p-tau217 and NfL [15, 16, 14]. A hypothesis-generating AD proxy score constructed from these features achieved AUC = 0.699 as a standalone synthetic-mortality predictor. This motivates a prospective cohort study measuring these biomarkers alongside directly assayed p-tau217 and NfL; it does not constitute a clinical finding and is presented as a direction for future investigation only.

### Limitations

Three constraints are worth stating directly. First, features were sampled independently; real EHR data have strong inter-feature correlations (for example, between albumin and creatinine in hepatorenal syndrome) that will shift importance rankings and potentially the ablation profile. Second, the performance gap over SOFA in the simulation is partly an artefact of the generating mechanism and will be smaller on real data where the true outcome function is unknown, covariates are correlated, and treatment decisions confound the mortality signal. Third, near-demographic-parity in the simulation is partly a construction artefact; real-world fairness evaluation on EHR data is necessary. CatBoost was replaced by HistGradientBoosting due to a NumPy 2.x binary incompatibility; this is a minor limitation as both models achieve comparable performance on tabular data.

## 6. Conclusions

We present and simulate-validate a complete pipeline for uncertainty-aware clinical triage: conformal prediction with formal distribution-free coverage guarantees, MC-Dropout epistemic–aleatoric decomposition, selective prediction with principled abstention, and a three-zone routing system. The pipeline was validated on a SOFA-calibrated synthetic ICU cohort (*N* = 90,000) calibrated to published epidemiology.

The key simulation-stage results are: conformal coverage verified at all tested miscoverage levels; UNCERTAIN zone AUC 0.624 at chance level, confirming correct routing of ambiguous cases; temporal AUC range 0.003 across four chronological windows; sex and age fairness gaps of 0.005 each. The novel lactate/albumin composite *r*_LA_, constructed from clinical first principles, ranks fifth among 32 features with a cross-seed CV below 1%.

The complete simulation confirms each component is correctly implemented and behaves as theoretically expected before contact with real patient data. External validation on MIMIC-IV — re-evaluating feature importance, conformal calibration, and fairness profiles on real patients — is the planned and necessary next step. All code and cohort generation scripts will be deposited on Zenodo upon acceptance.

## Supporting information

Supplementary Materia

## Data Availability

The primary dataset used in this study was a synthetic SOFA-calibrated ICU cohort generated for methodological evaluation. Additional publicly available datasets, including the UCI Heart Disease Dataset, Pima Indians Diabetes Dataset, and Stroke Prediction Dataset, were used exclusively for cross-dataset generalization testing. All data required to reproduce the analyses are either described within the manuscript or available from publicly accessible repositories. Further details regarding data generation and preprocessing are provided in the Methods section.

https://archive.ics.uci.edu/dataset/45/heart+disease

https://www.kaggle.com/datasets/uciml/pima-indians-diabetes-database

https://www.kaggle.com/datasets/fedesoriano/stroke-prediction-dataset

## Declarations

## Preprint disclosure

A preprint version of this work was posted to medRxiv (doi:10.64898/2026.05.29.26354474) prior to journal submission. The present manuscript is substantially revised relative to the preprint in framing, title, and scope; the underlying dataset and quantitative results are unchanged.

## Data and code availability

All Jupyter notebooks, cohort generation scripts, trained model weights, and result tables will be deposited on Zenodo upon acceptance. The three cross-domain datasets used in Supplementary Table S1 (Heart Disease UCI, Stroke, PIMA Diabetes) are publicly available from the UCI Machine Learning Repository [28].

## Ethics statement

No ethical approval was required for this study. The primary analysis used a fully synthetic cohort generated entirely from published epidemiological parameters; no human participants, biological samples, or patient records of any kind were involved at any stage. Cross-domain testing used three publicly available, de-identified benchmark datasets (Heart Disease UCI, Stroke, PIMA Diabetes) obtained from the UCI Machine Learning Repository. No protected health information was accessed. All procedures comply with relevant institutional guidelines at Dibrugarh University.

## CRediT authorship

**Amit Kalita:** Conceptualisation, Methodology, Software, Investigation, Formal analysis, Writing—original draft, Writing—review and editing. **Avirup Chattopadhyay:** Investigation, Data curation, Writing—review and editing. **Madhushree Bhattacharjee:** Validation, Formal analysis, Writing—review and editing. **Kaushik Das:** Supervision, Writing—review and editing.

## Declaration of AI-assisted technologies

During manuscript preparation, the authors used Paperpal (Cactus Communications) for language editing and grammatical refinement. The authors reviewed and edited all output and take full responsibility for the published content.

