## Supplementary Materia for "A Simulation-Based Validation Framework for Uncertainty-Aware Clinical Triage: Conformal Prediction and Ensemble Learning Applied to a Synthetic ICU Benchmark Cohort"

medRxiv 2026.05.29.26354474 (Version 2)

---

#### Contents

**Supplementary Note S1: ICU Biomarker and AD Pathway Overlap** **2**

**Supplementary Table S1: Cross-Domain Generalisation** **4**

### Supplementary Note S1: ICU Biomarker and AD Pathway Overlap — A Hypothesis for Future Study

**Motivation.** The features that dominate the mortality signal in this simulation — albumin, lactate, creatinine, BUN, and GCS — are also established confounders of the two leading plasma Alzheimer's disease (AD) biomarkers, phosphorylated tau-217 (p-tau217) and neurofilament light chain (NfL). Albumin alters p-tau217 protein binding [1]; renal dysfunction reduces p-tau217 clearance [2]; and NfL is significantly elevated in sepsis and correlates with lactate [3]. The partial dependence plot inflection points for these features (main paper, Fig. 9) align with thresholds from three independent clinical literatures: lactate 2.0 mmol/L (Sepsis-3 [4]), albumin 3.0 g/dL (clinical hypoalbuminaemia cutoff), and GCS below 13 (the 2024 Alzheimer's Association staging criteria [5]).

**AD proxy score construction.** To characterise this overlap quantitatively, a weighted composite AD proxy score was constructed from the eight features most mechanistically linked to p-tau217 or NfL axes (albumin, lactate, creatinine, BUN, GCS, gcs_sofa_ix, lactate_albumin_ratio, anemia_age_ix), with weights proportional to their permutation importance ranks. The proxy score was evaluated as a standalone predictor of synthetic mortality and compared against XGBoost probability.

**Results.** The proxy score achieved AUC = 0.699 as a standalone predictor of synthetic mortality. Its correlation with XGBoost probability was $r = 0.339$ — a correlation that is partly a mathematical consequence of shared feature ancestry between the proxy score and the XGBoost training set, and does not constitute independent evidence for an AD biomarker pathway. Stratifying by tertile yielded ICU mortality rates of 28.6%, 54.1%, and 67.3% (Fig. 1).

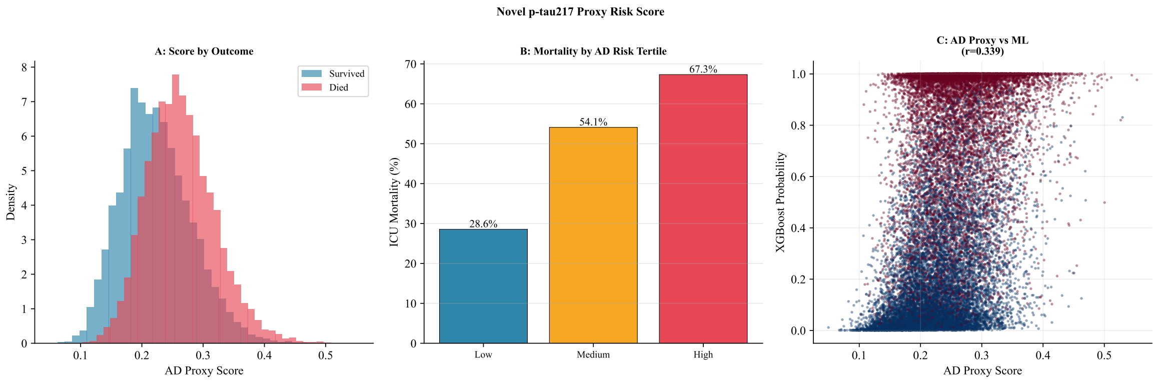


Figure 1: Hypothesis-generating AD proxy risk score (synthetic simulation). Panel A: score distributions by outcome (survived vs. died). Panel B: ICU mortality rate by tertile (28.6% → 54.1% → 67.3%). Panel C: correlation with XGBoost probability ($r = 0.339$); this reflects shared feature ancestry between the two scores and motivates prospective biomarker measurement, not a clinical finding.

**Interpretation and limitations.** These results are hypothesis-generating only. The proxy score was constructed and evaluated entirely within the synthetic simulation environment; its predictive signal is a mathematical consequence of the generating equation, which assigns large mortality coefficients to the same features the proxy score comprises. The $r = 0.339$ correlation with XGBoost probability reflects this shared ancestry and provides no independent evidence for a mechanistic AD biomarker pathway in real patients.

These observations motivate a prospective cohort study in which routine ICU biomarkers are measured alongside directly assayed p-tau217 and NfL in real patients. The computational proxy score built from synthetic data cannot serve as a substitute for that prospective measurement. This secondary analysis is included here as a hypothesis-generating supplement to the main pipeline validation and does not form part of the primary contribution of the paper.

### Supplementary Table S1: Cross-Domain Generalisation

Table 1 reports performance on three independent publicly available datasets via proxy feature mapping. These datasets played no role in model training and were used solely to test whether the acute organ-stress signal learned on the synthetic ICU cohort partially transfers to related or unrelated clinical domains.

Table 1: Cross-domain generalisation via proxy feature mapping. The pipeline was trained exclusively on the synthetic ICU cohort; these three datasets were used for post-hoc domain transfer testing only. Diabetes PIMA below-chance AUC confirms domain specificity: a model built on acute organ-failure physiology has no reason to generalise to population-level metabolic risk, and the failure is expected and informative.

| **Dataset** | *N* | **AUC** | **95% CI** | **Interpretation** |
|---|---|---|---|---|
| Heart Disease UCI | 920 | 0.703 | 0.666–0.735 | Partial transfer of acute organ-stress signal |
| Stroke | 5,110 | 0.748 | 0.716–0.778 | Partial transfer to cerebrovascular domain |
| Diabetes PIMA | 768 | 0.479 | 0.436–0.520 | Domain mismatch (expected; confirms specificity) |

All three datasets are publicly available and de-identified, obtained from the UCI Machine Learning Repository [6]. Proxy feature mapping was used to align available features with the 32 variables used during ICU training. AUC reported on full dataset with 1,000-resample bootstrap 95% CI.

**Interpretation.** The partial transfer results for Heart Disease UCI (AUC 0.703) and Stroke (AUC 0.748) indicate that acute physiological stress markers — shared across cardiovascular and cerebrovascular critical illness — carry some cross-domain discriminative signal. The below-chance PIMA Diabetes result (AUC 0.479) is the important control: it confirms the pipeline is learning acute organ-failure physiology, not a generic illness or demographic pattern that would spuriously generalise. The cross-domain results are exploratory and do not affect the primary conclusions of the paper.
